# Distinct pancreatic and neuronal Lung Carcinoid molecular subtypes revealed by integrative omic analysis

**DOI:** 10.1101/2021.07.27.21260865

**Authors:** Clara Domingo-Sabugo, Saffron A.G. Willis-Owen, Amit Mandal, Anca Nastase, Sarah Dwyer, Cecilia Brambilla, José Héctor Gálvez, Qinwei Zhuang, Sanjay Popat, Robert Eveleigh, Markus Munter, Eric Lim, Andrew G. Nicholson, Mark Lathrop, William O.C. Cookson, Miriam F. Moffatt

## Abstract

Lung Carcinoids (L-CDs) are uncommon low-grade neuroendocrine tumours that are only recently becoming characterised at the molecular level. Notably data on the molecular events that precipitate altered gene expression programmes are very limited. Here we have identified two discrete L-CD subtypes from transcriptomic and whole-genome DNA methylation data, and comprehensively defined their molecular profiles using Whole-Exome Sequencing (WES) and Single Nucleotide Polymorphism (SNP) genotyping. Subtype (Group) 1 features upregulation of neuronal markers (L-CD-NeU) and is characterised by focal spindle cell morphology, peripheral location (71%), high mutational load (*P*=3.4×10^−4^), recurrent copy number alterations and is enriched for Atypical Lung Carcinoids. Group 2 (L-CD-PanC) are centrally located and feature upregulation of pancreatic and metabolic pathway genes concordant with promoter hypomethylation of beta cell and genes related to insulin secretion (*P*<1×10^−6^). L-CD-NeU tumours harbour mutations in chromatin remodelling and in SWI/SNF complex members, while L-CD-PanC tumours show aflatoxin mutational signatures and significant DNA methylation loss genome-wide, particularly enriched in repetitive elements (*P*<2.2 × 10^−16^). Our findings provide novel insights into the distinct mechanisms of epigenetic dysregulation in these lung malignancies, potentially opening new avenues for biomarker selection and treatment in L-CD patients.

## Introduction

Lung carcinoids (L-CDs) are rare, slow growing neuroendocrine tumours that represent 2% of all lung cancers^1^ and 30% of well-differentiated Neuroendocrine Tumours (NETs) throughout the body^2^. Diagnosis is currently based on criteria in the WHO Classification of Thoracic Tumours^3^, namely a combination of neuroendocrine morphology, mitotic index and presence or absence of necrosis, supported by immunohistochemistry for neuroendocrine markers.

The latest World Health Organization (WHO) classifies L-CDs as part of the spectrum of neuroendocrine neoplasms (NEN), with typical and atypical L-CDs grouped under the term NETs, and Small Cell Lung Cancer (SCLC) and Large Cell Neuroendocrine Carcinoma (LCNEC) under the term Neuroendocrine Carcinomas (NECs), more in keeping with other organ systems^3,4^. NETs frequently occur in never-smokers, in contrast to NECs. ACs are more aggressive than TCs, with greater likelihood of metastases at presentation and poorer survival. Thus, TCs and ACs are considered low grade and intermediate grade respectively, while LCNECs and SCLC are classified as high-grade. These groupings based on histopathology have been supported by recent genomic and mutational studies that have shown similar genes to be altered across NENs, but differences between NECs and NETs, and within SCLC, LCNEC and NETs themselves, although at different prevalences between NETs, with the lowest number of molecular alterations in L-CDs^5–9^.

Lung carcinoids remain relatively understudied despite their increasing incidence^10–16^. Although many patients with rare NETs can be cured by resection, management of advanced disease remains problematic^17^, and clinically experience no clear pathway of care in their cancer journey, with their tumours difficult to diagnose and treat and patients lacking disease specific support^18^. Disagreement exists over the optimal diagnosis^2^ and there is inter-observer variability in differentiating TCs from ACs^19^.

To date no lung carcinoid studies have investigated DNA methylation alterations at a genomic scale or have integrated genetic, transcriptomic and clinical data. We therefore describe here a comprehensive characterization of these malignancies at the molecular level.

## Results

### Molecular classification from transcriptomic data

Unsupervised clustering and Principal Components Analysis (PCA) analysis of RNA sequencing data from 15 LCAR tumours (comprising 3 ACs and 12 TCs) clearly differentiated tumours into two approximately equally sized groups (n=8 and 7 respectively). These two groups were reproducibly identified using data from the top 500 most variable genes or all sequenced genes (*n* 25,764) (Supplementary Fig. 1a-b; Supplementary Table 1). Substantial differential expression was present between these groups, with 1,924 transcripts achieving significance at a 1% False Discovery Rate (FDR) threshold (Fig. 1; Supplementary Data 1). Table 1 lists the top 20 transcripts.

**Figure 1:**
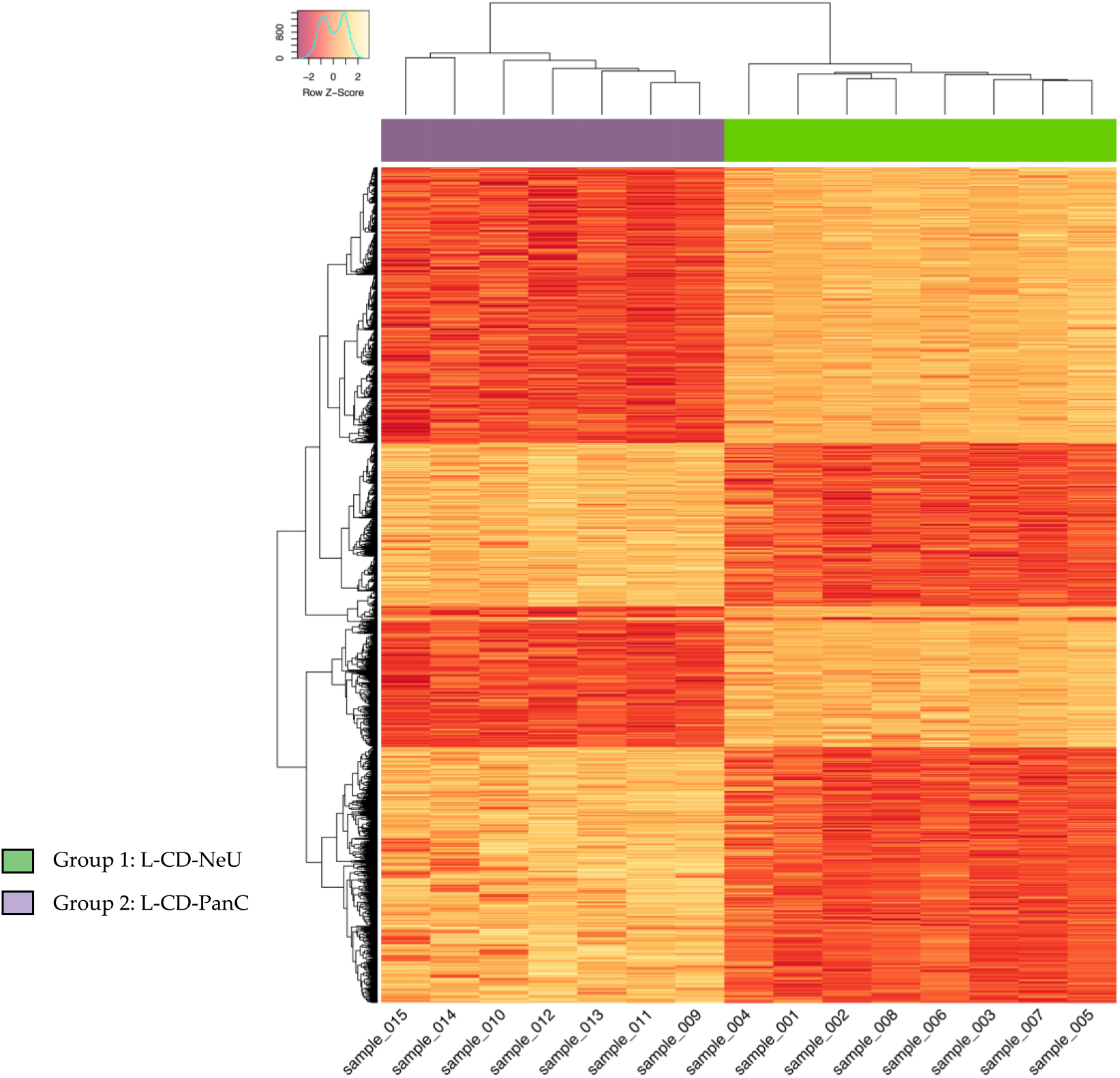
Heatmap of the significantly differentially expressed (DE) genes (*adj*.*P*<0.01) between L-CD subtypes. Figure displays heat map and dendrograms with hierarchical clustering of 15 lung carcinoids (on the X axis) and genes (on the Y axis) that were significantly differentially expressed. Top bar indicates lung carcinoid group membership.

**Table 1.** The top 20 transcripts differentially expressed between PanC and NeU L-CDs (*adj*.*P*<0.01).

| *Gene* | *Log<sub>2</sub>FC (CI)* | *AveExpr* | *P.Value* | *adj.P* | *PanC* | *NeU* |
| --- | --- | --- | --- | --- | --- | --- |
| *A1CF* | 10.63(6.53,14.73) | 3.24 | 2.94E-13 | 1.89E-09 | Red | Blue |
| *TM4SF5* | 10.50(10.25,10.73) | 1.46 | 2.02E-14 | 3.39E-10 |  |  |
| *SERPINA10* | 9.77(8.37,11.17) | 1.12 | 4.39E-12 | 1.03E-08 |  |  |
| *RDH12* | 9.65(8.00,11.30) | 4.20 | 8.05E-13 | 3.69E-09 |  |  |
| *RFX6* | 9.42(8.65,10.20) | 1.44 | 2.12E-13 | 1.82E-09 |  |  |
| *HNF1A* | 8.33(8.04,8.63) | 1.16 | 2.63E-14 | 3.39E-10 |  |  |
| *C2orf72* | 7.30(6.79,7.81) | 3.41 | 2.02E-12 | 5.79E-09 |  |  |
| *FOXA3* | 7.24(6.91,7.58) | 0.60 | 8.18E-12 | 1.76E-08 |  |  |
| *ARHGEF10L* | 3.02(2.54,3.51) | 6.78 | 1.00E-12 | 3.69E-09 |  |  |
| *GLYCTK* | 2.91(2.21,3.62) | 4.82 | 1.18E-12 | 3.79E-09 |  |  |
| *RFC3* | -2.44(-4.40,-0.47) | 4.29 | 1.09E-08 | 3.65E-06 | Blue | Red |
| *AUTS2* | -2.51(-2.91,-2.11) | 4.99 | 1.51E-08 | 4.62E-06 |  |  |
| *ATP8A1* | -2.89(-3.67,-2.11) | 5.83 | 3.62E-09 | 1.56E-06 |  |  |
| *DPYSL3* | -5.08(-6.09,-4.06) | 7.31 | 1.10E-07 | 2.08E-05 |  |  |
| *DOK7* | -5.53(-5.99,-5.07) | -1.07 | 1.30E-08 | 4.23E-06 |  |  |
| *PRUNE2* | -5.58(-6.22,-4.94) | 4.63 | 4.71E-08 | 1.18E-05 |  |  |
| *FAM3B* | -6.21(-7.12,-5.29) | 0.98 | 8.76E-08 | 1.76E-05 |  |  |
| *HS3ST6* | -6.27(-6.71,-5.82) | -0.39 | 2.82E-08 | 7.66E-06 |  |  |
| *KCNK10* | -9.52(-9.86,-9.18) | 0.84 | 3.03E-09 | 1.40E-06 |  |  |
| *RALYL* | -9.70(-10.31,-9.08) | 0.21 | 1.73E-10 | 1.78E-07 |  |  |

Abbreviations: log<sub>2</sub>FC (log<sub>2</sub> fold change); CI (confidence interval); AveExpr (Average expression across all samples in log<sub>2</sub>-counts per million); adj.P (Benjamini-Hochberg false discovery rate adjusted *p*-value). Coloured cells indicate relatively higher (red) or lower (blue) gene expression between subtypes.

GSEA revealed significantly enriched metabolic pathways and hallmarks of pancreatic beta cells in Group 1 (Supplementary Fig. 2), peaking at regulation of beta cell development (Normalised Enrichment Score [NES] 2.19, *P*<0.01). Consistent with these themes, the genes showing the strongest evidence of differential expression, both with relatively raised expression in this group, were *TM4SF5* (log_2_FC 10.50, adj.*P*=3.39×10^−10^) and *HNF1A* (log_2_FC 8.33, adj.*P*=3.39×10^−10^). *TM4SF5* is a known tumorigenic factor in several cancer types, including liver, colon, pancreatic and esophageal cancers^20–25^, whilst *HNF1A* encodes a putative master regulator of human pancreatic cancer stem cell properties^26^. The APOBEC1 complementation factor *(A1CF)* showed the highest log_2_ fold change (log_2_FC) in expression as compared to Group 2, suggesting that A1CF protein expression could be used as a molecular marker.

Group 2 showed upregulation of pathways involved in neuronal differentiation, peaking at serotonin neurotransmitter release cycle (NES −1.79, *P*=0.004). In line with this pattern, we observed significantly higher expression levels of various neuronal genes in this group (Supplementary Data 1). *ASCL1* for example, encodes a neuronal differentiation transcription factor and is a lineage-specific oncogene for high-grade neuroendocrine lung cancer^27,28^, whilst *SLIT1, ROBO1* and *SRGAP1* all represent members of the cell signalling protein complex slit/robo which is involved in axon guidance and angiogenesis. In addition, FAM3B/PANDER, a pleiotropic secreted cytokine that induces apoptosis in insulin-secreting beta-cells^29^ was highly expressed in Group 2 (log_2_FC 6.21, adj.*P=*1.76×10^−5^). PANDER is expressed ubiquitously, including lung^30^ and some neurons of the brain^31^, and it has recognised roles in invasiveness and tumorigenicity when overexpressed in prostate^32^ and colon^33^ cancers.

On the basis of these features, from this point onwards we refer to Group 1 as LCAR-NeU and Group 2 as LCAR-PanC.

### L-CD subtypes are associated with histopathological parameters

Integration of our findings with clinical parameters showed that LCAR-NeU tumours were characterised by a focal spindle cell morphology (two-sided Fisher’s exact test: estimate – 17.50 [95% CI 1.24, 227.8], *P*=0.041) and peripheral location (two-sided Fisher’s exact test: estimate – 0 [95% CI 0, 0.46], *P*=0.007) (Figs. 2a and 2b) and included every tumour of an Atypical Lung Carcinoid histology (*n* 3). LCAR-PanCs on the other hand, were all located centrally. Whilst LCAR-NeU patients exhibited a trend towards an older age, this difference was not statistically significant (Welch’s *t*-test: t −-2.01; estimate PanC=53.63, estimate NeU=68.86 [95% CI −32.06, 1.59]; df 10.5; *P*=0.071). No significant association was seen with survival, smoking history, sex, presence of emphysema, nodal stage and either lymph or vascular invasion.

**Figure 2:**
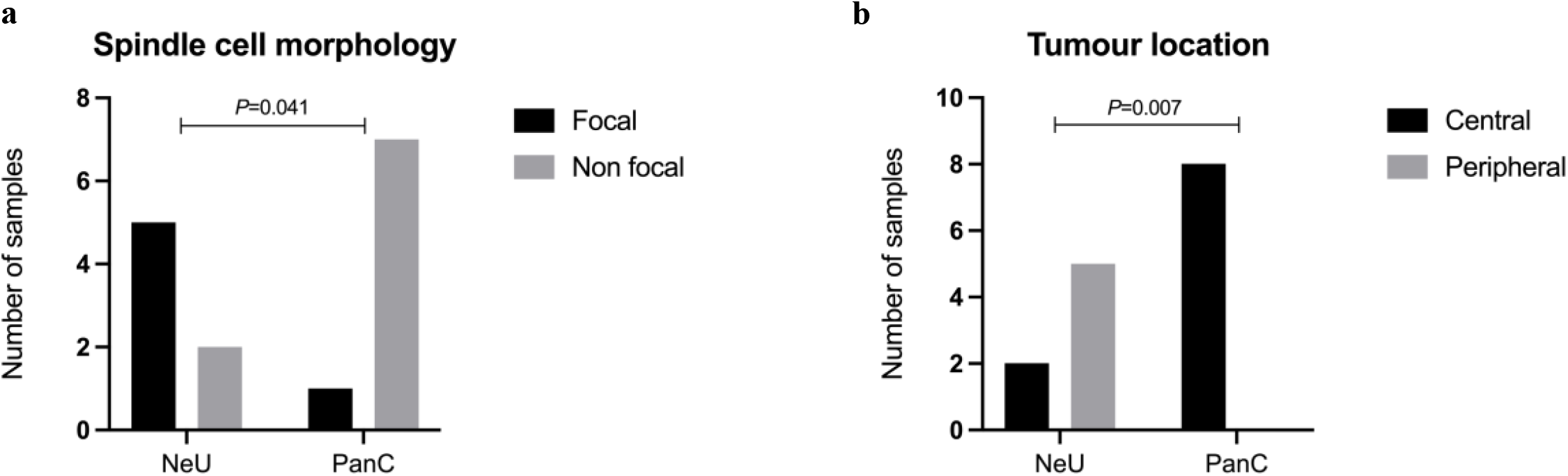
L-CD molecular groups have distinct histological characteristics. a) Focal spindle cell morphology in L-CD-NeU versus L-CD-PanC. b) Central or affecting the main lobar broncos versus peripheral tumour location between L-CD-NeU and L-CD-PanC. *P*-values are for two-sided Fisher’s statistical tests.

### Mutational signatures of L-CD subtypes

We sought to investigate the molecular features associated with these contrasting expression programmes by analysing the spectrum of somatic base substitutions and their trinucleotide context. The mutational load was significantly higher in LCAR-NeU with 45.83 mutations on average, compared with 27.87 for LCAR-PanC (two-sided *t*-test: t −4.66; estimate PanC=27.87, estimate NeU=45.83 [95% CI −26.36, −9.56]; df 12; *P=*5.53×10^−4^) (Supplementary Fig. 3a). This is consistent with a higher mean number of mutations detected in ACs, and tumours from this histology all falling in the LCAR-NeU molecular group.

A different spectrum of COSMIC Mutational Signatures (CMS) was consistently observed between the two LCAR groups; 50% of LCAR-PanC samples possessed an aflatoxin exposure signature. This is consistent with our differential gene expression data, in which several genes (*GGT5, CYP34A, CYP3A5* and *DPEP1*) participating in aflatoxin activation and detoxification were expressed at significantly higher levels in PanC tumours.

On the other hand, LCAR-NeU samples variably showed a CMS 5 signature (57%), which is found in most cancer samples, and/or a CMS 8 signature (43%), which is associated with double strand break repair by homologous recombination. A summary of the mutational signatures is given in Supplementary Figure 4a-b.

Since most observed signatures were of unknown aetiology, we identified the *de novo* mutational signatures and related these to catalogued (COSMIC) signatures^34^. Analysis with a cophenetic correlation metric confirmed a consistent presence of different spectra of mutational signatures in the two groups. LCAR-PanC showed an aflatoxin signature (CSM 24; cosine similarity of 0.603), while LCAR-NeU showed spontaneous deamination of 5-methylcytosine (CSM 1; cosine similarity of 0.801). Both groups were found to share a mutational signature associated with defective DNA mismatch repair (CSM 20; cosine similarities of 0.399 in PanC and 0.642 in NeU) (Supplementary Fig. 5). These data confirm a biological distinction between the two observed LCAR subtypes.

### Mutations and Copy Number Alterations (CNAs) in L-CD subtypes

The most frequently mutated genes in L-CD-NeU were components of the cytoskeleton (57%), including *ITGA1, ITGA2, PLEC* and *TPJ1*, and histone covalent modifiers (43%) (*JMJD1C, KDM1B* and *KDM4E*) (Fig. 3). By contrast, L-CD-PanC tumours showed mutations in *ARID1A* and *ARID5B* (25%), both members of the SWI/SNF complex; and the Notch signalling genes *PLXND1* and/or *WWC1* (25%). Notch signalling controls cell fate decisions and has previously been found to be activated in neuroendocrine cells undergoing reprogramming after injury^35^.

**Figure 3:**
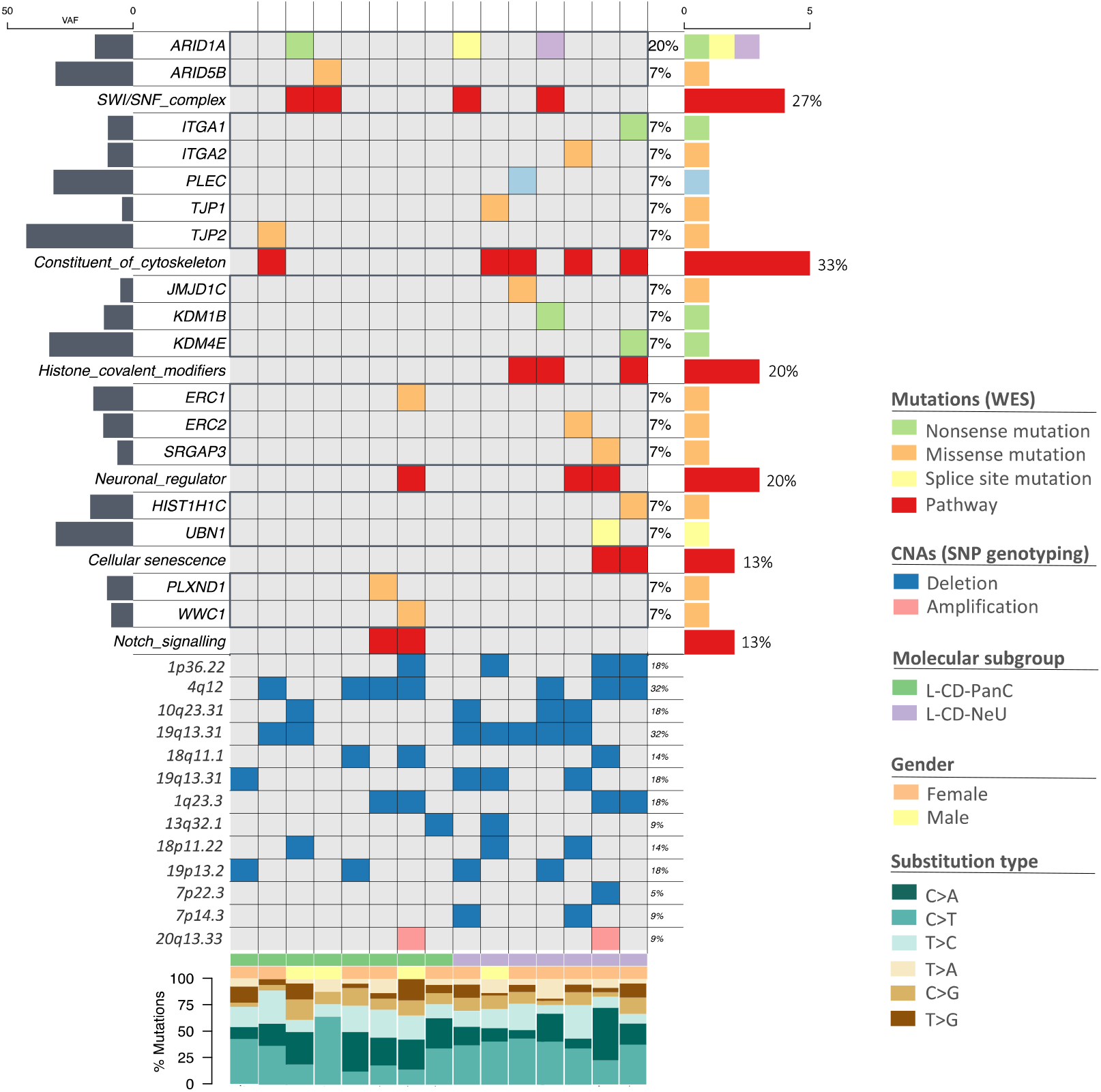
Mutational landscape of −LCD molecular groups. Oncoplot of the most recurrent mutations, InDels and significant copy number alterations (CNAs) in LCAR-NeUs (*n*=7) and LCAR-PanCs (*n*= 8). Abbreviations: VAF (Variant Allele Frequency); WES (Whole-Exome Sequencing).

No significant difference in copy number burden (CNB) could be detected between the two L-CD classifications. Just 4.8 and 5.5% of the genome showed evidence of CNB in L-CD-NeU and L-CD-PanC respectively (inter-sample range L-CD-NeU: 0.3%-14.5%; L-CD-PanC: 0.3%-19.3%). These values lie within the range previously defined in healthy human populations^36^.

Individual CNAs were found in genes related to the innate immune system and neutrophil degranulation, and included deletions affecting C1orf127 (1p36.22), *TXK* and *TEC* (4q12), *NDUFS2* (1q23.3), *KIF20B* (10q23), *INMT, ROCK1* and zinc finger proteins (*ZNF180, ZNF846, ZNF283, ZNF404*). One significant amplification was found in *SCYP2*, a major component of the synaptonemal complex during meiotic division. A summary of the genes harbouring mutations and CNAs is given in Figure 4 with information regarding genes in significant cytobands and the percentages for CNAs in each subtype are detailed in Supplementary Table 2.

**Figure 4:**
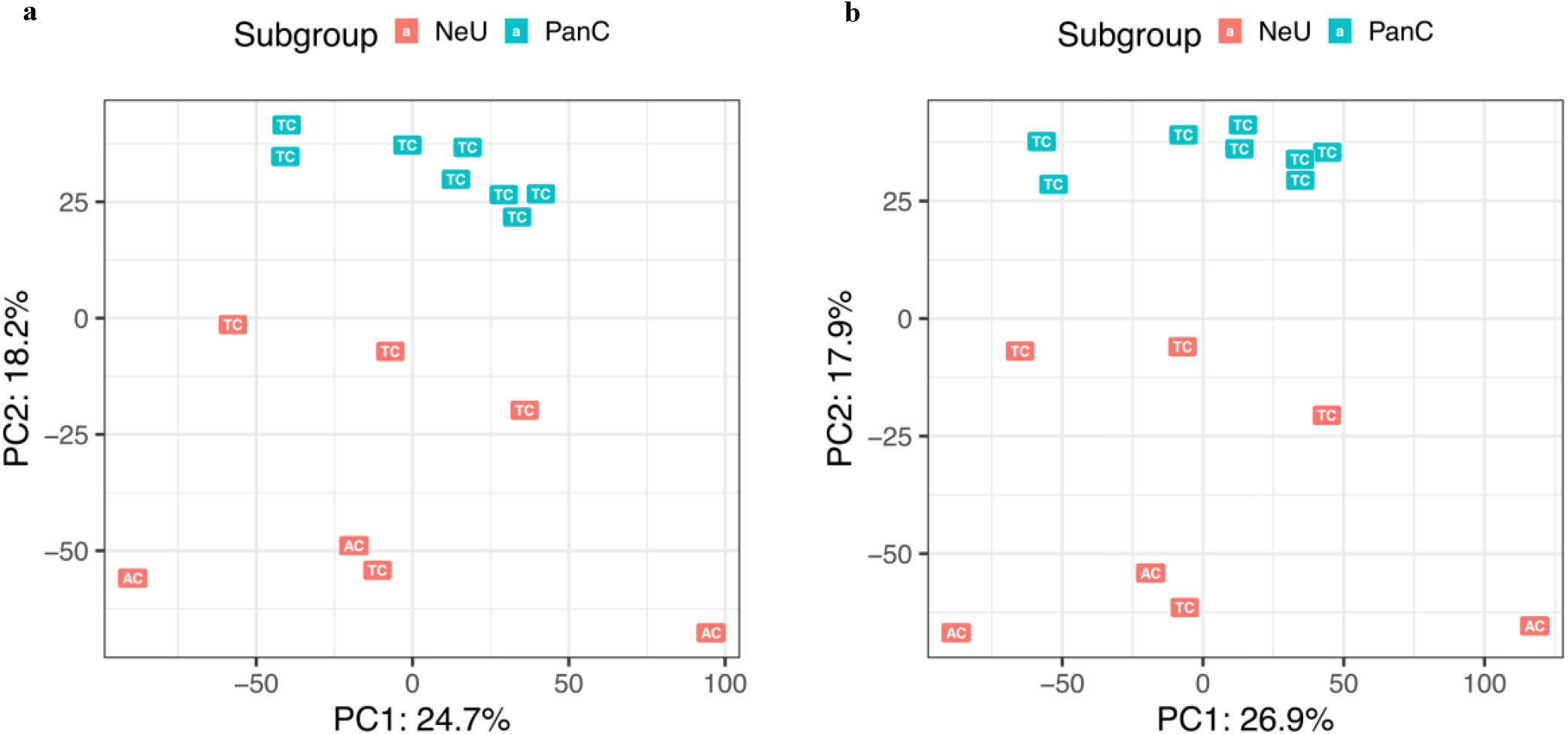
Principal components analysis of whole genome (a) and repeat element (b) CpG DNA methylation data differentiates L-CD molecular groups. Figure shows principal components analysis of WGBS data, a: whole genome, b: repeat elements only. NeU tumours are shown in red, PanC tumours are shown in blue. Histology is shown as TC and AC for Typical Carcinoid and Atypical Carcinoid, respectively.

Similar to mutations, L-CD-NeU showed higher frequencies of CNAs, with 71.43% of tumours harbouring *KIF20B* deletions, a kinesin involved in neuron polarization^37^. In contrast with earlier studies^38^, we did not detect any significant loss in *RB1, TP53* and *MEN1* or any significant gains in *TERT, SDHA* or *RICTOR*.

### Focal and widespread DNA methylation changes distinguish L-CD subtypes

Aberrant DNA methylation is a common feature of cancer and provides a mechanism of gene expression dysregulation. Here we applied whole genome bisulfite sequencing (WGBS) to generate a snapshot of the DNA methylation profile in tumour and matched histologically normal tissue. Mirroring our observations from transcriptomic data, PCA of genome-wide DNA methylation data differentiated L-CD-NeU from L-CD-PanC tumours (Fig. 4a). We identified 4,304 significant Differentially Methylated Regions (DMRs) distinguishing these L-CD groups (Supplementary Data 2), and consistent with gene expression data, we identified promoters of genes expressed in beta cells and related to insulin secretion as hypomethylated in the L-CD-PanC group (Fig. 5). Other hypomethylated genes included *SMAD7, NRG1* and *PMS1*. On the other hand, L-CD-NeU tumours showed hypomethylation of the PI3K/AKT/mTOR signalling pathway, a pathway known to be involved in pluripotency and cell fate determination, and *HOXB2* and *HOXB* developmental genes were also found to be hypomethylated. *HOX* genes are major transcriptional regulators. They have key roles in development, and frequent epigenetic and/or transcriptional deregulation in cancer ^39,40^.

**Figure 5:**
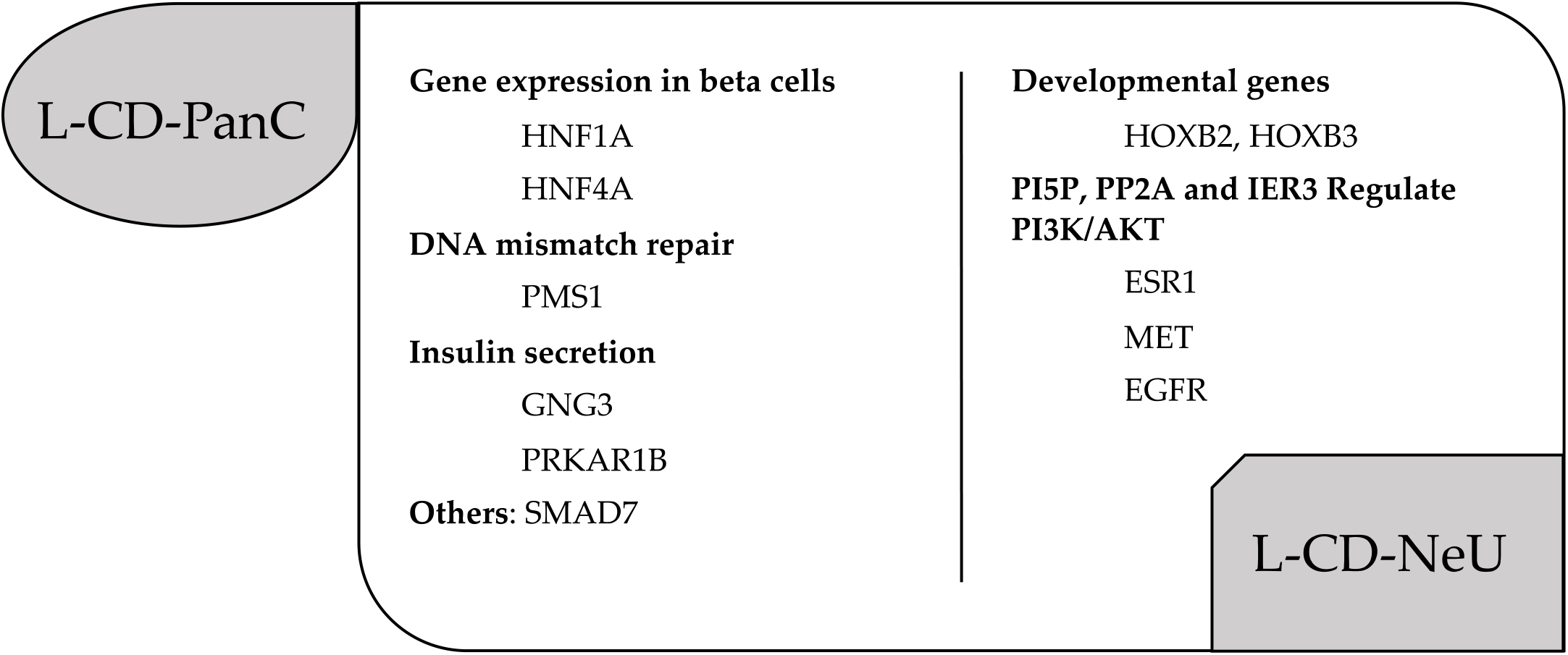
Genes with promoters showing significant differential methylation between L-CD molecular groups. Statistical analysis was performed using a Wald test and *P*<1×10^−6^ was considered statistically significant. Only DMRs with a methylation difference of >20% between the two groups were considered for annotation.

Nevertheless, most DMRs mapped to intergenic and intronic regions (Supplementary Table 3). We therefore sought to investigate the properties of DNA methylation alterations in the non-coding genome of the two L-CD groups. PCA analysis of different genomic regions’ DNA methylation levels revealed that repeated elements explained most of the variance (26.89%) and alone distinguished L-CD-NeU from L-CD-PanC (Fig. 4b).

Reactivation of transposable elements (TE) through epigenetic mechanisms is an established feature of some cancers with roles in tumour immunity^41^. We found here a significant enrichment of TEs in hypomethylated DMRs (relative enrichment = 1.31, 95% CI 1.16-1.48, *P*=8.03×10^−06^). Specifically, DMRs enriched in TEs with a 30% fraction overlap were significantly lowly methylated (two-sided Wilcoxon matched-pairs signed rank test: V 446264, estimate 0.03 [95% CI −0.04, −0.03], *P*<2.2 × 10^−16^) in PanC tumours (Supplementary Fig. 6). Repeat elements and non-genic CpG sites are known to lose methylation during aging^42^, nevertheless, we found no significant difference in age between LCAR subtypes.

## Discussion

In this study we have identified two distinct molecular subtypes of L-CDs from RNA-sequencing data; L-CD-NeU and L-CD-PanC, and characterised their molecular profiles using WES, SNP genotyping and WGBS. We have shown that these two groups differ significantly in their transcriptional, mutational and epigenetic profiles, as well as their physical characteristics.

Differential analysis of gene expression data showed upregulation of metabolic pathways and hallmarks of pancreatic beta cells in PanC tumours, whereas pathways related to the neuronal system, neurotransmitter synthesis and release were found enriched in NeU tumours. Differential expression between these two groups was most marked with *TM4SF5*, notable since TM4SF5-targeted monoclonal antibody and peptide vaccination has previously established preventive and/or therapeutic effects in hepatocellular carcinoma, colon cancer and pancreatic cancer models^25^. On the basis of these data, assessment of anti-hTM4SF5 antibody treatment efficacy in TM4SF5-expressing PanC L-CDs may be warranted.

In our transcriptomic data, *A1CF* showed the highest log_2_ fold-change expression between the two L-CD groups suggesting that A1CF protein expression may have utility as a molecular marker for anti-hTM4SF5 antibody therapy. Furthermore, we show that the NeU L-CD classification is significantly associated with a focal spindle cell morphology and peripheral tumour location, meaning that these physical features may provide a minimally invasive, accessible proxy, in agreement with the distinct characteristics of central and peripheral carcinoids reported by George et al^43^. Additionally, FAM3B/PANDER expression has been detected at the protein level and its inhibition has shown antitumour effects *in vitro* in prostate and several human cancer lines^30^, suggesting therapeutic potential and a marker for the differential diagnosis of L-CD subtypes in combination with TM4SF5 and A1CF protein expression.

Molecular profiling of L-CDs have previously shown chromatin remodelling genes, such as *MEN1, ARID1A, PSIP1, KMT2C* and *KMT2A*, to be recurrently mutated in L-CDs while *TP53, RB1* and *STK11* mutations have been found frequently altered in non-carcinoid NETs^44^. Other studies have emphasized the distinction between TCs and ACs, and molecular events distinguishing these subtypes are reported to affect the genes *MEN1, TP53, KMT2C, TERT, SDHA, RICTOR* and *RB1*^6^. In our study we did not detect any mutation or CNA in any of these genes, in contrast with earlier studies^6,28^.

L-CD-NeU showed a higher tumour mutational load affecting cytoskeletal genes and histone covalent modifiers. Conversely, L-CD-PanC tumours showed a lower mutational load, mostly affecting members of the SWI/SNF complex and Notch signalling pathways. L-CD-NeU tumours also showed more recurrent CNAs than L-CD-PanCs. Although L-CD-NeUs encompassed all the ACs, most members of this group had typical histology, highlighting the potential importance of molecular screening to help in the therapy decision process.

To gain insights into the biological mechanisms involved in L-CD carcinogenesis, we looked for the most frequent combinations of somatic mutations and identified a combination of known and *de novo* mutational signatures. Known mutational signatures from the COSMIC database (https://cancer.sanger.ac.uk/signatures/) revealed a mixed repertoire of signatures that have been found in other cancer types. Consequently we identified *de novo* signatures and compared them with known catalogued COSMIC signatures. Both approaches linked an aflatoxin signature with L-CD-PanC, whereas signatures previously found in all cancer types were identified in the L-CD-NeU group.

Aflatoxin B1 (AFB1) is a potent genotoxin produced by species of the *Aspergillus* fungus. It can bind to double stranded DNA^45^ and induce hepatocellular carcinoma leaving a C→A mutational signature^46,47,48^. Pancreatic tumours have previously showed dominance of this signature potentially due to the mutational properties of AFB1-DNA adducts^49,50,51,52^. Extrahepatic tissues, such as the nasal olfactory and respiratory mucosa, and mucosa of the trachea and oesophagus have a high capacity to activate AFB1 which when inhaled may cause lung cancer^53,54^. Here we observed predominance of C→A mutations in the L-CD-PanC group (Supplementary Fig. 3b). High levels of AFB1 can be present in respiratory grain-dust particles^55,56^, and as such could potentially contribute to L-CD-PanC carcinogenesis. Controlled experiments designed to detect and define any role of aflatoxin in L-CD-PanC pathogenesis should be a priority for future investigation.

The mutational landscapes of L-CDs alone could not explain the transcriptomic differences detected and so we analysed their DNA methylomes by using WGBS data. We identified DMRs in promoters of pancreatic beta cells and genes related to insulin secretion in L-CD-PanCs, as well as mismatch repair genes, pinpointing DNA methylation changes as a key event in this cancer group.

TE-enriched regions showed significant hypomethylation in L-CD-PanCs and alone were sufficient to differentiate from L-CD-Neu tumours by PCA analysis. This suggests that epigenetic dysregulation in the non-coding genome may be a major contributor to the PanC subtype. Significantly higher expression levels of *A1CF* in L-CD-PanC tumour genomes may contribute to this generalised hypomethylation. A1CF serves as a docking site to recruit APOBEC1 deaminase, which has a role in active DNA demethylation followed by T:G mismatch repair^57,58^. Altogether these findings highlight the value of whole-genome data to better understand and refine the molecular alterations in these malignancies.

We recognise a number of limitations of our study. The relative rarity of L-CD tumours and access to fresh frozen tissue has meant that our sample size is fairly small. Histopathological analyses with the markers we have identified could validate the observed classifications and should be a priority for future studies to enable translation into a clinical setting.

The distinct clinical phenotypes associated with these molecular subtypes suggest novel biomarkers for patient stratification. Our findings could be used to refine and complement the current WHO classification of lung carcinoids and will stimulate further investigation to improve the cancer journey of patients with L-CD

## Data Availability

Genotyping and sequencing data generated during the current study will be submitted at the European Genome-Phenome Archive (EGA) (https://ega-archive.org/) under accession code: EGAS00001005473.

## Methods

Paired tumour and adjacent normal tissue samples were obtained from 15 patients with a carcinoid diagnosis who had undergone tumour resection at the Royal Brompton Hospital between year 2010 and year 2014. Samples were collected prior to therapy and snap frozen at the time of surgical resection for genomic analyses. For transcriptomics, tissue was stored within two hours in RNAlater. Methods for genomic DNA and RNA extraction have been described previously^59^. Patient consent and tissue banking was carried out under appropriate ethical approval (RBH NIHR BRU Advanced Lung Disease Biobank [NRES reference 10/H0504/9] and Brompton and Harefield NHS Trust Diagnostic Tissue Bank [NRES reference 10/H0504/29]). Tumour diagnosis using the WHO classification and tumour cell abundance were determined through pathology review (AG Nicholson) of haematoxylin and eosin staining. Clinical data is summarised in Supplementary Table 4.

### RNA-sequencing (RNA-Seq) and Gene Set Enrichment Analysis (GSEA)

Total RNA was isolated with the RNEasy Fibrous Midi Kit (Qiagen, Hilden, Germany) from 15 tumour samples. Samples were analysed with the 2100 Bioanalyser and total RNA Nano Kit (Agilent Technologies, California, United States) following manufacturer’s instructions. The Illumina TruSeq stranded total RNA Gold sample preparation protocol (RS-122-2301) was used to prepare libraries. RNA sequencing was performed at the Imperial BSC Genomics Facility using the HiSeq4000 system with sequencing by synthesis (SBS) chemistry for paired ends. Raw sequencing reads were quality and adapter trimmed using cutadapt (v.1.9.1) in Trim Galore. Reads were aligned against the December 2013 Human Genome Assembly (GRCh38/ hg38) using STAR (v. 2.7.3a). Processing of the gene count data was performed using voom as implemented in the limma package (v. 3.42.2). Normalised log2-counts per million were used as a measure of gene expression. A heatmap of RNA expression was produced using Euclidean distance across samples with the heatmap.2() function in gplots package (ver. 3.1.1). Clustering validation measures were obtained with clValid R package (ver. 0.7). Differential Expression (DE) analysis was modelled using limma (ver. 3.42.2) and the empirical Bayes method. *P*-values were adjusted using a Benjamini Hochberg method to control the FDR below 1%.

The Gene Set Enrichment Analysis (GSEA) (ver. 4.0.3) software was utilised to detect significant group-wise differences in a Reactome Canonical Pathways Gene Set^60^. In the setting of exploratory discovery, we used a less conservative FDR of 0.25 and nominal *P*-value of 0.01.

### WES and mutational analysis

WES and mutation calling was performed at the McGill Genome Centre, Montreal, Canada. Sequencing libraries were prepared with the SureSelect^XT^ Target Enrichment System (Agilent SureSelect Human All Exon V4) and sequenced with 100bp Paired-End Illumina HiSeq2000 Sequencer.

Processing and variant calling of the WES data was performed using the analysis piepeline “GenPipes”^61^. In short, Fastq files were trimmed and aligned against the Human Genome December 2013 Assembly (GRCh37/hg19) with BWA mem. SAM files thus obtained per sample were sorted by chromosomic location with GATK (Genome Analysis Tool Kit) (ver.3.7) and read alignments deemed to be PCR duplicates were removed with Picard (ver. 2.9.0). Pre-processing involved local realignment around known insertions/deletions (InDels) and Base Quality Score Recalibration (BQSR) with GATK (ver. 3.7). Somatic mutations and InDels were called with MuTect (ver. 1.16) and Scalpel (ver. 0.4.1) software, respectively. Somatic calls were then combined, and subsequent steps involved decomposing multiallelic variants from VFC files, genetic variant annotation and functional effect prediction with SnpEff (ver. 4.3) and addition of metadata with Genome MINIng (GEMINI) (ver. 0.14-0.20) software. The annotated variants were filtered based on known impact predictions and CADD scores. We considered pathogenic somatic variants as those with predicted high or medium impact, and those assigned a minimum CADD score of 15 and not flagged as polymorphic.

### Identification of Mutational signatures

The R (ver.3.6.1) DeconstructSigs package (ver. 1.8.0) was used to determine the contribution of known mutational processes in L-CD samples. Signature matrices were calculated based on the fraction of times a mutation was seen in each of the 96-trinucleotide context for each COSMIC catalogued signature (v2). The exome2genome normalisation method was used to reflect the absolute frequency of each nucleotide context as it would across the whole genome. As a result, a reconstructed mutational profile was obtained based on the final weights for each individual sample.

### SNP genotyping and identification of Copy Number Alterations (CNAs)

Illumina Infinium OmniExpressExome (v1.6) arrays were used for genotyping of > 900,000 SNP markers in 15 tumour and normal matched L-CD samples. The processing and Quality Control were performed with GenomeStudio (ver. 2.0.4). SNPs in pseudo-autosomic regions and InDels were filtered out.

To improve accuracy of copy number variant detection ASCAT (v.2.5.2) was separately applied in tumour and normal samples for GC bias correction. DNACopy (v.1.56.0) was used to segment copy number data into regions of estimated equal copy number. Regions with >10 markers support were used for calling CNAs with Gistic2 (ver. 2.0.23). Germline Copy Number Variants (CNVs) were first identified on normal lung tissue samples and were subtracted from the tumour segmented data before CNA calling. Copy number data was visualised using Maftools (ver. 1.9.30).

### Whole Genome Bisulfite Sequencing (WGBS-Seq) and Differential Methylation analysis

WGBS libraries were constructed using the KAPA High Throughput Library Preparation Kit (Roche/KAPA Biosystems) from 1ug of the sperm DNA spiked with 0.1% (w/w) unmethylated lambda and pUC19 DNA (Promega). DNA was sonicated (Covaris) and fragments sizes of 300-400bp were controlled on a Bioanalyzer DNA 1000 Chip (Agilent). Following fragmentation, DNA end repair of double stranded DNA breaks, 3’-end adenylation, adaptor ligation and clean-up steps were conducted according to KAPA Biosystems’ protocols. The sample was then bisulfite converted using the Epitect Fast DNA bisulfte kit (Qiagen) following manufacturer’s protocol. The resulting bisulfite DNA was quantified with OliGreen (Life Technology) and amplified with 9-12 PCR cycles using the KAPA HiFi HotStart Uracil+ DNA Polymerase (Roche/KAPA Biosystems) according to suggested protocols. The final WGBS libraries were purified using Ampure Beads, validated on Bioanalyzer High Sensitivity DNA Chips (Agilent) and quantified by PicoGreen (ThermoFisher). Sequencing was carried out with the paired-end Illumina HiSeq X Next Generation Sequencing System at the McGill Genome Centre, Montreal, Canada. Analysis of WGBS-Seq data was performed using the Methyl-Seq GenPipes adapted from the Bismark pipeline^61^. Alignment of paired-end reads to bisulfite converted Human Genome (GRCh37/hg19) was performed with bismark (ver. 0.18.1) and bowtie2 (ver. 2.3.1) according to the bismark user guide using the non-directional option. GATK (ver. 3.7) and Picard (ver. 2.9.0) were used to sort by chromosomal location and for the removal of PCR duplicates. The bismark methylation extractor was used to extract methylation in CpG context. Bis-SNP was used to identify SNPs and insertion/deletion events (InDels) that were subsequently filtered out with Bedops (ver. 2.4.35).

Normalisation of the processed WGBS data was carried out with the methylKit R package (ver. 1.12.0). Finally, Dispersion Shrinkage for Sequencing data with single replicates (DSS-single) implemented in the DSS Bioconductor R package (ver. 2.34.0) was used for calling of Differentially Methylated Regions (DMRs) to account for spatial correlation, read depth and biological variation between groups. Specifically, a smoothing span of 500 bps was used to improve estimation of methylation levels by accounting for nearby CpG sites. A Wald statistical test was then performed for CpGs with at least 10% difference between tumour and normal, and the difference was considered significant when *P*-value <1×10^−6^. Regions a minimum of 50 bps in length with at least 3 significant CpGs were identified as DMRs and merged when co-localised by less than 50 bps. Finally, only DMRs with an absolute tumour:normal methylation difference exceeding 20% were selected for further analysis. DMRs were annotated into different categories with the annotatr R package (ver. 1.12.1).

## Acknowledgements

We thank the patients for participating to our study. We also thank Dr. Alain Pacis and Paul Stretenowich for their help with the analysis of WGBS data. This study was supported by the Asmarley Trust and by the Wellcome Trust. Sample collection for this study was supported by the NIHR Respiratory Disease Biomedical Research Unit at the Royal Brompton and Harefield NHS Foundation Trust. The funding sources had no role in study design, collection, analysis and interpretation of the data, or in the writing of the manuscript.

## Author contributions

WOCC and MFM designed and conceptualised the study. EL, AGN and CB recruited patients with histological assessment and meta data provided by CB and AGN. SD performed the RNA sequencing. ML and MM were responsible for the WES and WGBS sequencing with ML directing generation of the data. CDS prepared the SNP genotyping libraries and associated data analysis with input from AN and AM. CD, RE and ML performed the somatic mutation variant calling. CDS performed the somatic InDel calling, DNA methylation and transcriptomic data analysis with input from AM, SW-O, JHG and QZ. SP provided critical input on the findings. CDS wrote the manuscript with input from SW-O, WOCC and MFM.

## Data availability

Genotyping and sequencing data generated will be submitted at the EGA European Genome-Phenome Archive (EGA) (https://ega-archive.org/) under accession code: EGAS00001005473.

## Additional Information

Supplementary Material is available for this paper.

