## Supplementary Material for "Distinct pancreatic and neuronal Lung Carcinoid molecular subtypes revealed by integrative omic analysis"

**Supplementary Figure 1. (a) Principal Component Analysis and (b) Dendrogram of sample similarity based on all sequenced genes (n=25,764).**

Abbreviations: TC (Typical Carcinoid; AC (Atypical Carcinoid); PC (Principal Component).

**a**

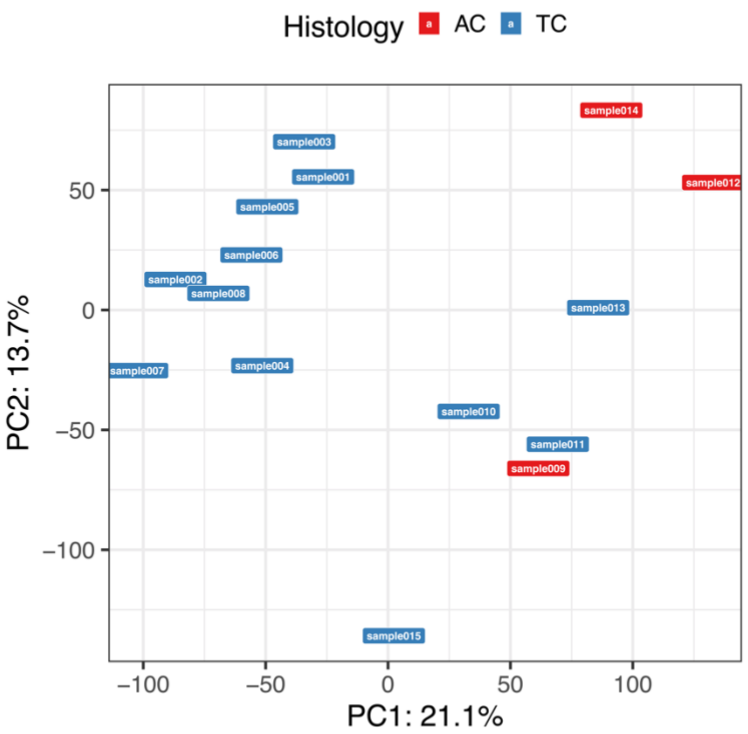


**b**

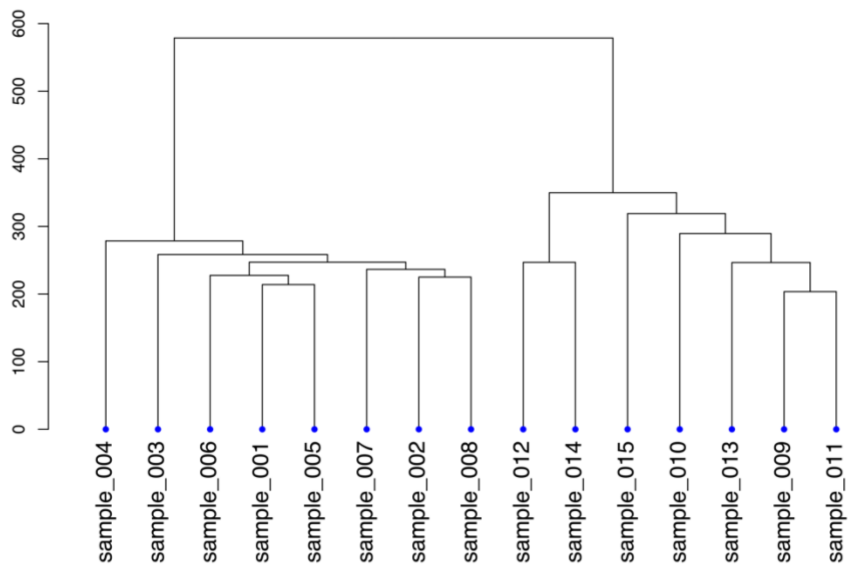

**Supplementary Figure 2: Reactome pathways nominally enriched in L-CD molecular subtypes.**

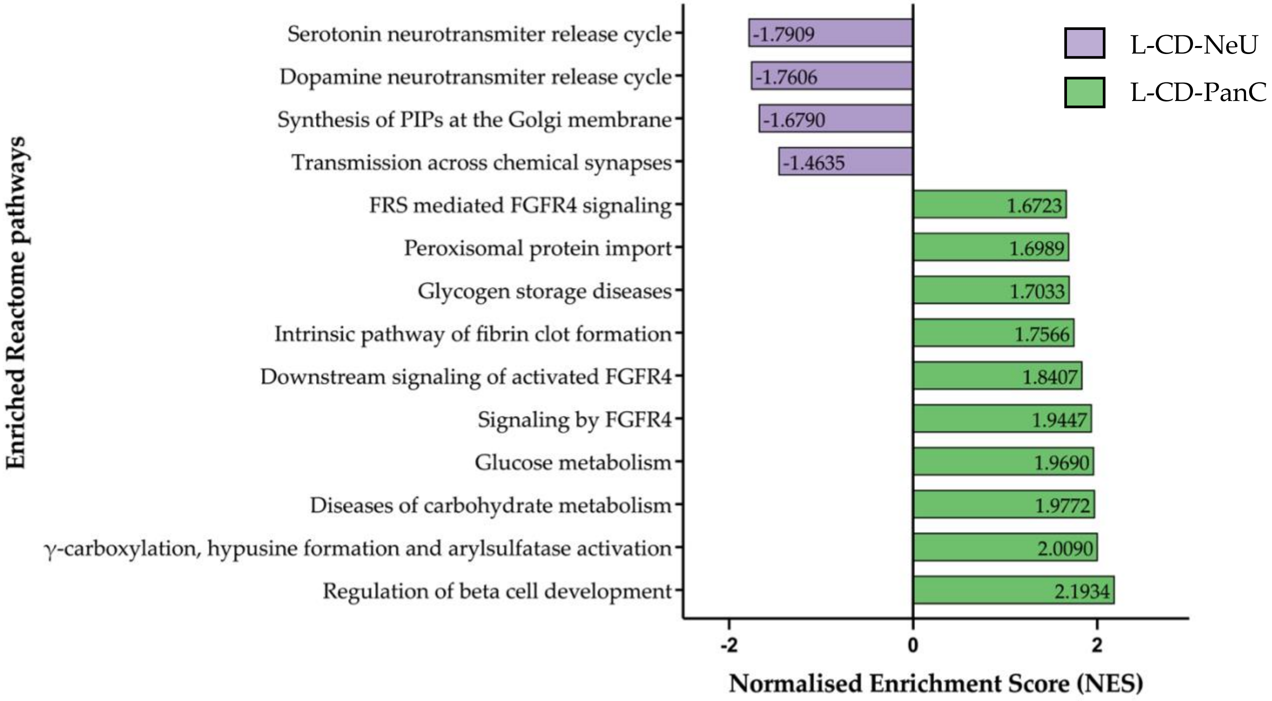


Bar plot showing the normalised enrichment scores of Reactome pathways achieving nominal significance for enrichment in a Gene Set Enrichment Analysis (GSEA) of RNA-sequencing data at a *p*-value threshold of 0.01. Using the MSigDB Reactome Canonical Pathways gene set[57], we find 10 enriched and 4 depleted pathways in the L-CD-PanC subtype relative to the L-CD-NeU subtype. Each bar represents a pathway and their Normalised Enriched Scores (NES) is given.

**Supplementary Figure 3: Mutational patterns of L-CD subtypes. (a)** Number of mutations in L-CD molecular subgroups was significantly higher in the L-CD-NeU subgroup (two-sided *t*-test, $P$=5.53x$10^{-4}$). wel(b**)** Types of substitutions. L-CD-PanC was characterised by a higher amount of C>A substitutions, whereas L-CD-NeU showed a higher number of mutations with a predominance of C>T substitutions, although differences were not statistically significant (Unpaired *t*-test: $P_{C>A}$=0.72; $P_{C>T}$=0.34).

**a**

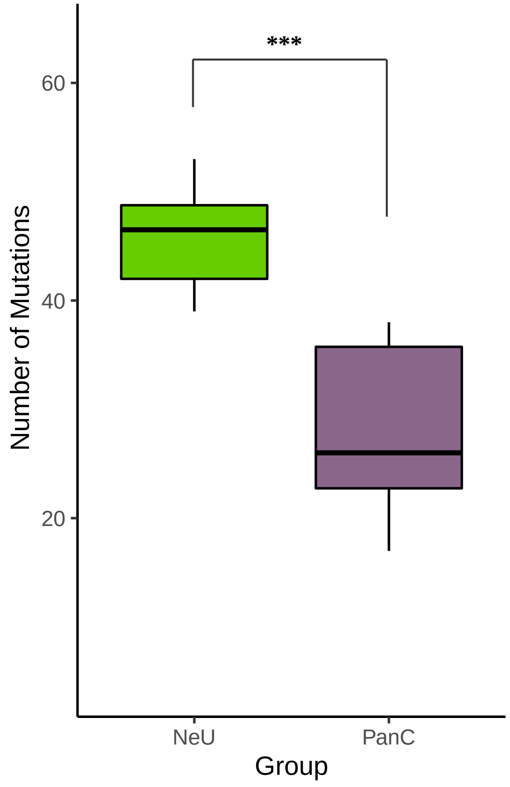


**b**

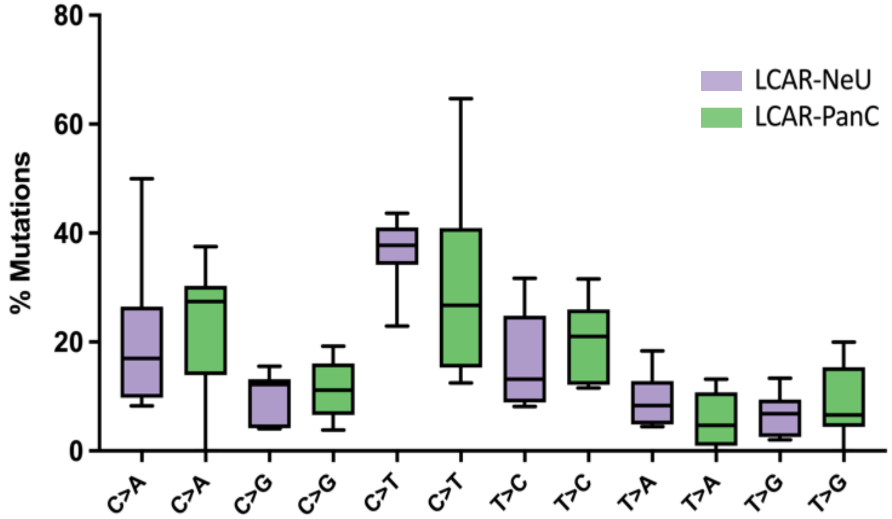

**Supplementary Figure 4: Weights of each mutational signature operative in (a) L-CD-PanC and (b) L-CD-NeU tumours.** Mutational Signatures identified with deconstructSigs R package in each molecular group with COSMIC mutational signatures version 2 by using the exome2genome normalization method.

a) **L-CD-PanC**

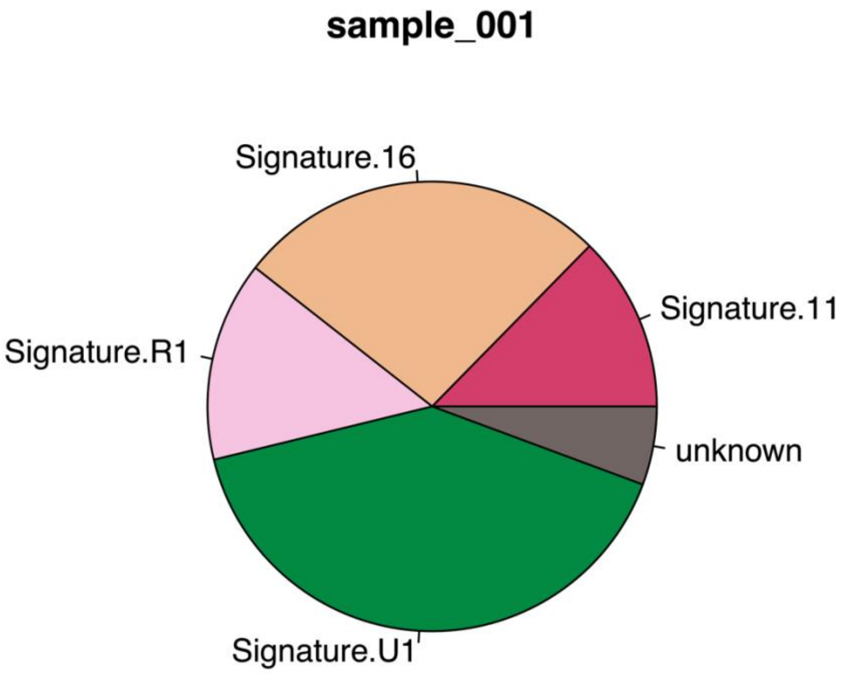

**sample_002**

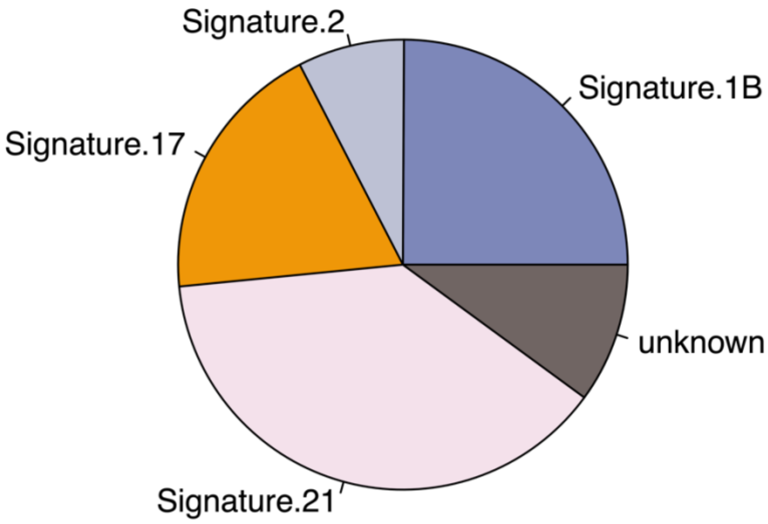


**sample_003**

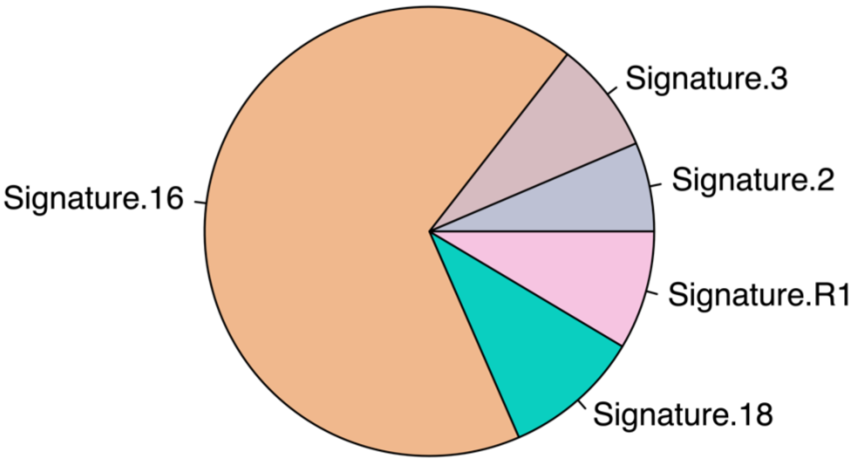

**sample_004**

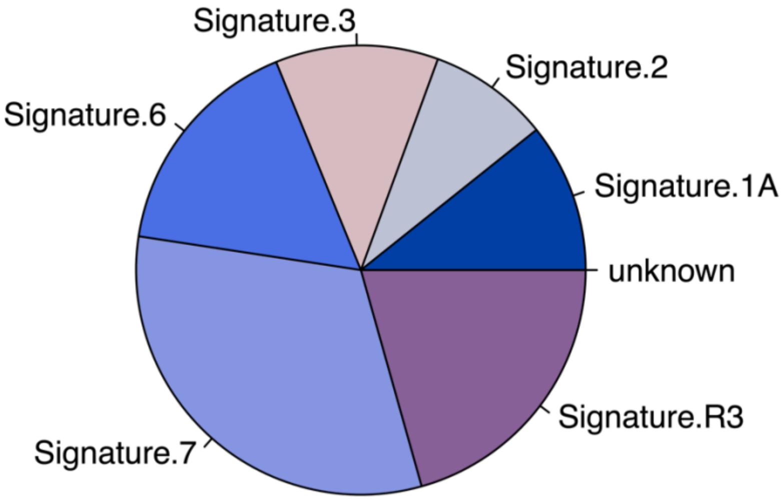


**sample_005**

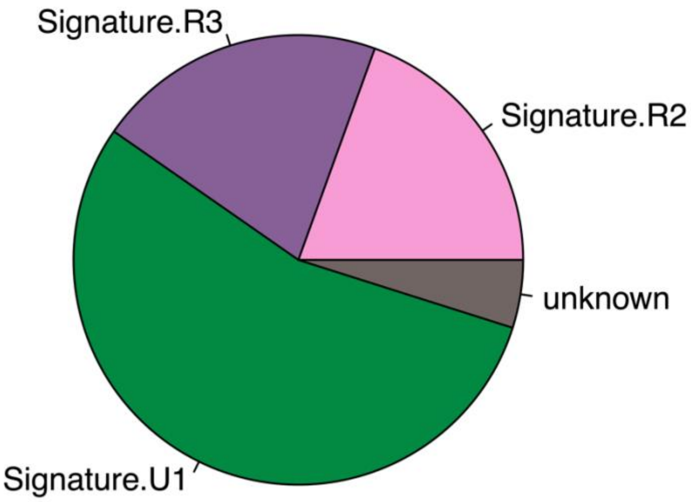

**sample_006**

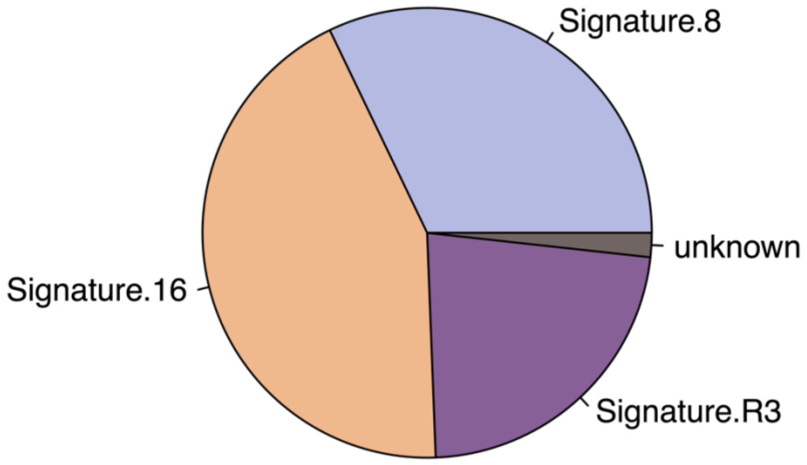


**sample_007**

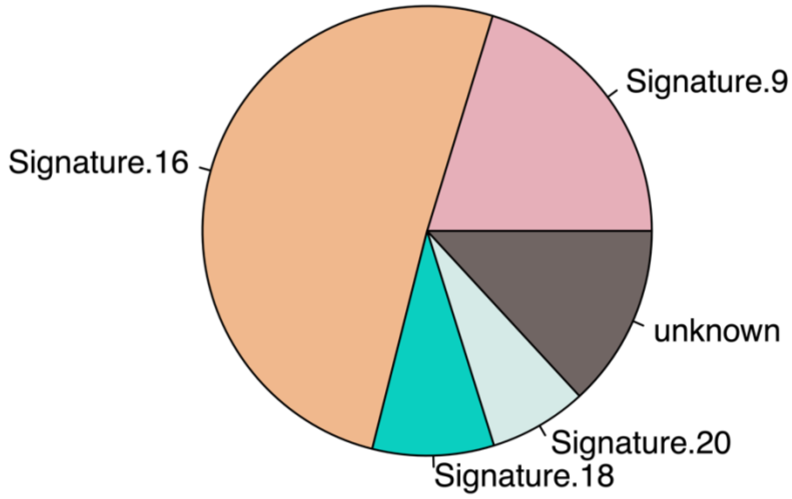

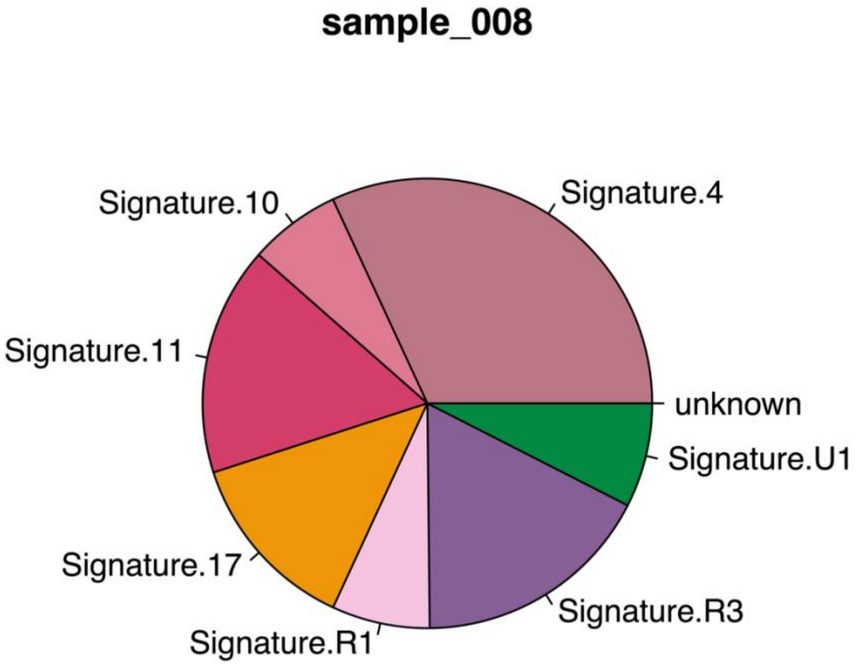
sample_008
Signature.4
Signature.10
Signature.11
unknown
Signature.U1
Signature.17
Signature.R3
Signature.R1

**b) L-CD-NeU**

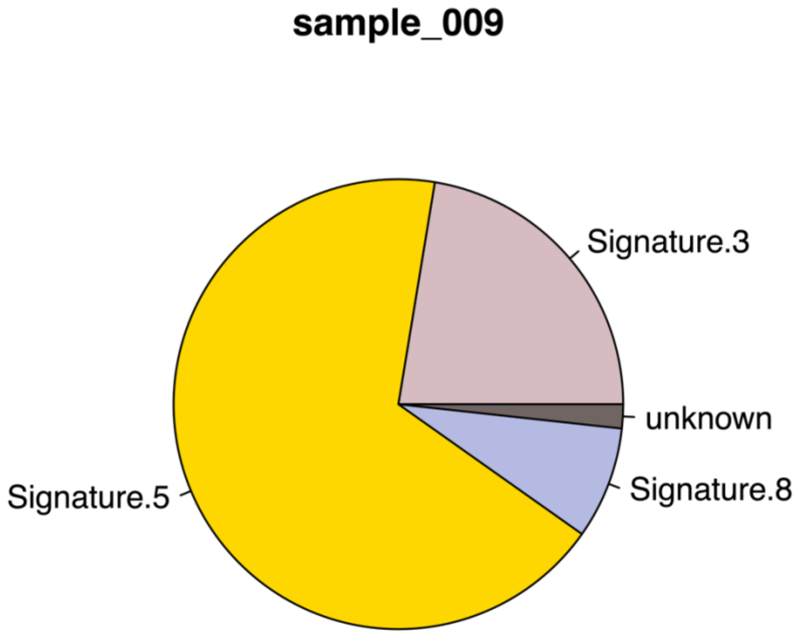
sample_009
Signature.3
unknown
Signature.5
Signature.8

**sample_010**

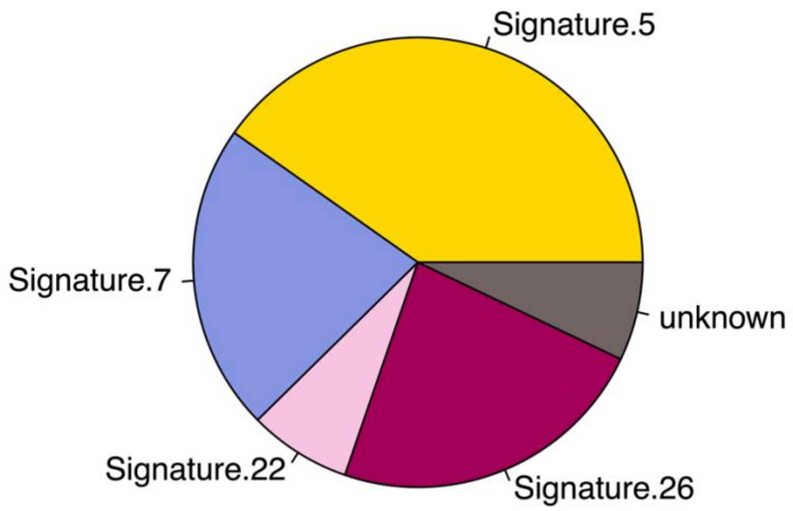


**sample_011**

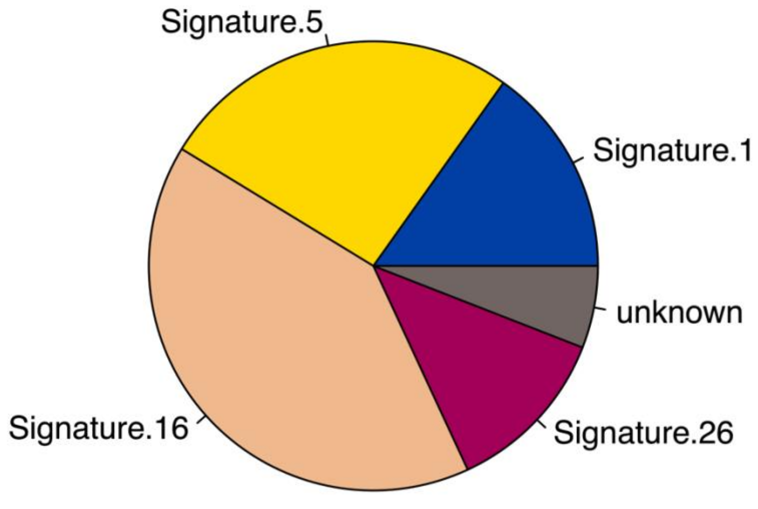

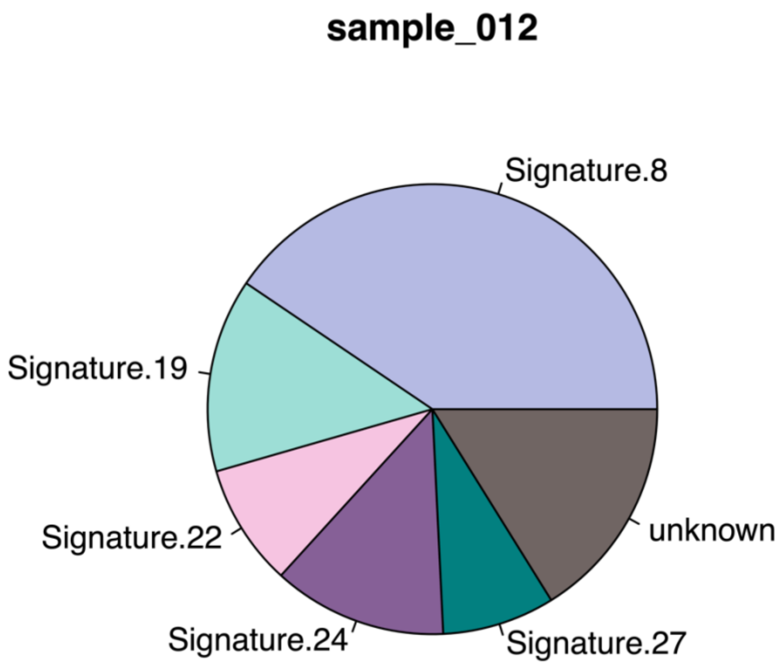
sample_012
Signature.8
Signature.19
Signature.22
Signature.24
Signature.27
unknown

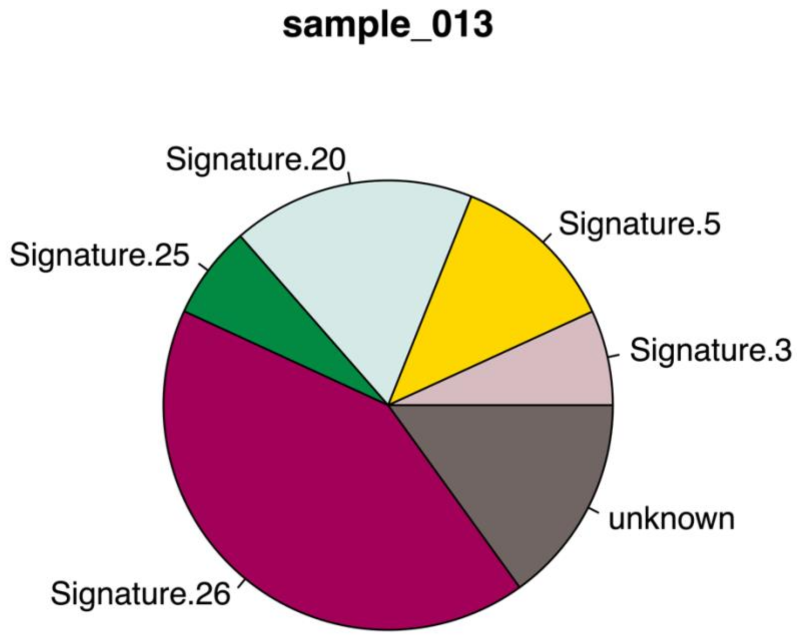
sample_013
Signature.20
Signature.5
Signature.25
Signature.3
unknown
Signature.26

**sample_014**

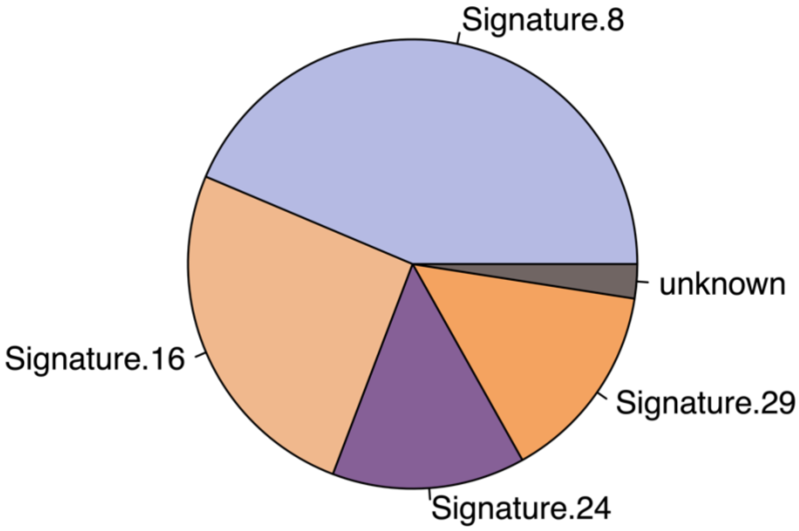


**sample_015**

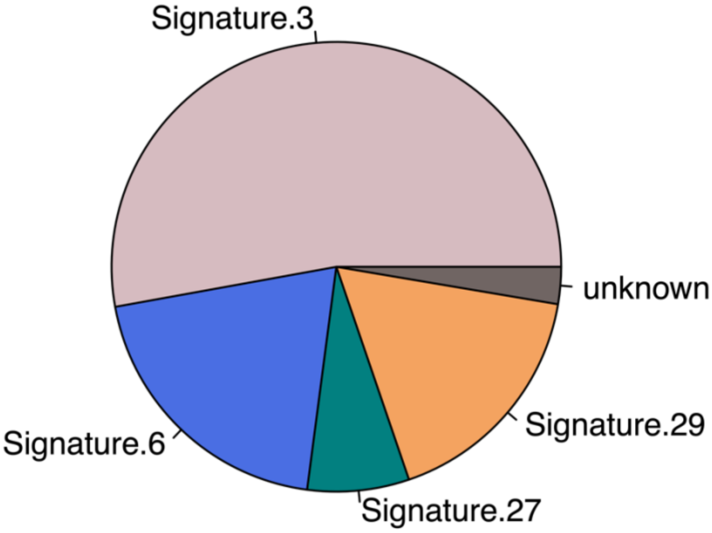

**Supplementary Figure 5:** ***De novo*** **Mutational signatures identified in L-CD molecular groups using matrix factorization and compared to known mutagenic processes (COSMIC mutational signatures) identified by Alexandrov and colleagues[49].** Abbreviations: CMS, Mutational Signature.

L-CD-PanC

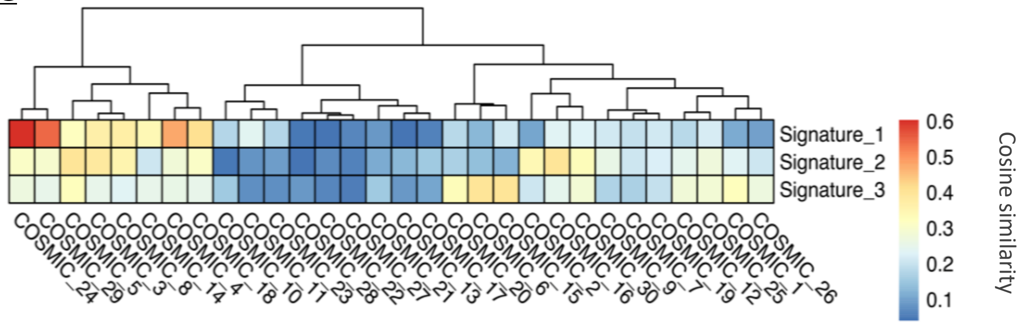


CMS 24 associated to exposure to aflatoxin (C>A)

L-CD-NeU

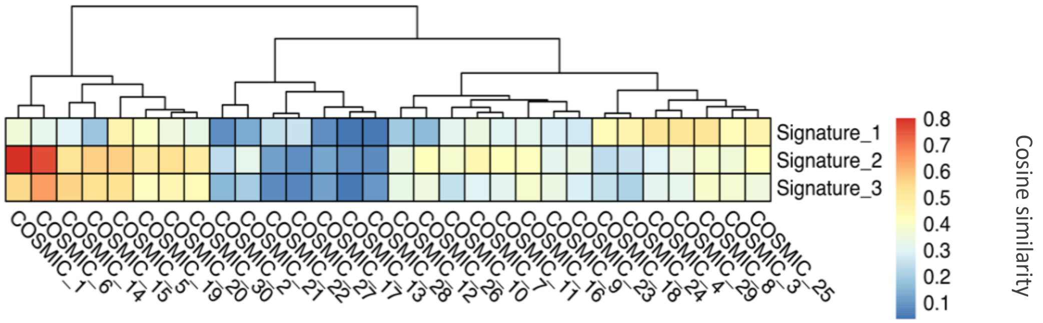


CMS 1 arising from spontaneous deamination of 5-methyl cytosine (C>T)

Common CMS 20 associated with defective DNA mismatch repair.

**Supplementary Figure 6: Boxplot of mean CpG DNA methylation levels in DMRs enriched with Transposable Elements (TEs).** L-CD-PanC tumours showed significant hypomethylation at TEs overlapping DMRs (two-sided Wilcoxon matched-pairs signed rank test: V 446264, estimate 0.03 [95% CI -0.04, -0.03], $P<2.2 \times 10^{-16}$). DMRs with an overlap fraction of >30% of the DMR length were only considered.

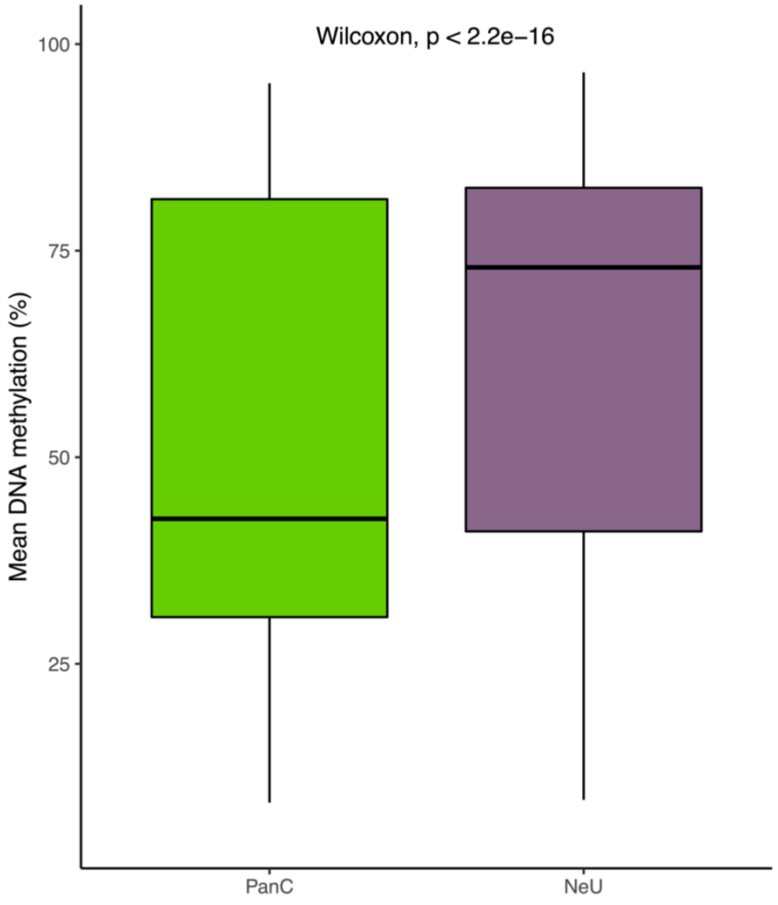

**Supplementary Table 1: Hierarchical clustering validation measures and optimal number of clusters identified using top 500 most variable genes with clValid R Package.** The optimal number of clusters using the hierarchical clustering algorithm was 2, thus validating the results obtained with hclust and PCA methods.

| MEASURE | SCORE | METHOD | OPTIMAL NUMBER OF CLUSTERS |
|---|---|---|---|
| Connectivity | 6.0833 | hierarchical | 2 |
| Dunn | 0.9540 | hierarchical | 2 |
| Silhouette | 0.3639 | hierarchical | 2 |

**Supplementary Table 2: Genes in significant cytobands (residual q-value <0.05) identified with GISTIC2 in L-CD molecular groups**.

| *Cytoband* | *Wide Peak Limits* | *Genes* | *q values* | *Residual q values* | *PanC (%)* | *NeU (%)* |
|---|---|---|---|---|---|---|
| ***1p36.22*** | chr1:11005496-11008843 | *C1orf127* | 4.28E-12 | 4.28E-12 | 12.50 | 42.86 |
| ***4q12*** | chr4:48068784-52743948 | *TXK, TEC* | 2.01E-09 | 2.01E-09 | 50.00 | 42.86 |
| ***1q23.3*** | chr1:161167699-161176136 | *ADAMTS4, NDUFS2* | 1.15E-07 | 1.15E-07 | 12.50 | 42.86 |
| ***10q23.31*** | chr10:91497196-91511197 | *KIF20B* | 1.07E-05 | 1.07E-05 | 25.00 | 71.43 |
| ***19q13.31*** | chr19:44978299-44988638 | *ZNF180* | 1.60E-05 | 0.00035745 | 25.00 | 14.29 |
| ***7p14.3*** | chr7:30793498-30795447 | *INMT* | 0.00042248 | 0.0038469 | 12.50 | 42.86 |
| ***18q11.1*** | chr18:14585295-18904233 | *ROCK1, ANKRD30B* | 0.00099715 | 0.00099715 | 25.00 | 28.57 |
| ***20q13.33*** | chr20:58416429-58467096 | *SCYP2* | 0.0073662 | 0.0073662 | 12.50 | 14.29 |
| ***19p13.2*** | chr19:9868450-9872906 | *ZNF846* | 0.0084492 | 0.0084492 | 12.50 | 28.57 |
| ***19q13.31*** | chr19:44350719-44377800 | *ZNF283, ZNF404* | 0.0084492 | 0.22424 | 25.00 | 28.57 |
| ***13q32.1*** | chr13:96496859-96546879 | *UGGT2* | 0.011732 | 0.011732 | 0.00 | 14.29 |
| ***7p22.3*** | chr7:2748718-2752382 | *AMZ1* | 0.017476 | 0.12272 | 0.00 | 28.57 |
| ***18p11.22*** | chr18:9254407-9256298 | *ANKRD12* | 0.017476 | 0.017476 | 12.50 | 14.29 |

Q-values for each called peak and the associated residual q-values after removing segments shared with higher peaks are shown. Percentage of samples harbouring each CNA is given for each molecular subtype.

**Supplementary Table 3: Number of DMRs between L-CD-PanC and L-CD-NeU per annotation type identified with annotatr R package.**

| *Annotation type* | *DMRs (n)* |
|---|---|
| *hg19_cpg_inter* | 3869 |
| *hg19_cpg_islands* | 60 |
| *hg19_cpg_shelves* | 194 |
| *hg19_cpg_shores* | 291 |
| *hg19_enhancers_fantom* | 174 |
| *hg19_genes_1to5kb* | 292 |
| *hg19_genes_3UTRs* | 89 |
| *hg19_genes_5UTRs* | 86 |
| *hg19_genes_exons* | 390 |
| *hg19_genes_introns* | 2265 |
| *hg19_genes_promoters* | 129 |
| *hg19_lncrna_gencode* | 497 |

Abbreviations: DMR (Differentially Methylated Region).

**Supplementary Table 4. Clinical data for L-CD patients.**

| *Sample ID* | *Gender (f/m)* | *Age range* | *Stage* | *Deceased (T/F)* | *Survival (months)* | *Smoking category* | *Tumour percentage* | *BMI* | *Molecular group* | *Histology* | *Location* | *Spindle cell morphology* | *Lymph invasion* | *Emphysema presence* |
|---|---|---|---|---|---|---|---|---|---|---|---|---|---|---|
| *sample_001* | f | *>71* | IB | F | 71 | NS | | 28 | PanC | TC | Central | No | No | Yes |
| *sample_002* | f | *>71* | IIB | F | 69 | NS | | 33 | PanC | TC | Central | Prevalent | Yes | Yes |
| *sample_003* | m | *51-70* | IA2 | F | 66 | ES | 80 | 26 | PanC | TC | Central | No | No | No |
| *sample_004* | m | *51-70* | IB | F | 61 | ES | 90 | 33 | PanC | TC | Central | No | Yes | No |
| *sample_005* | f | *25-50* | IA | F | 77 | NS | | 25 | PanC | TC | Central | Prevalent | No | Yes |
| *sample_006* | f | *51-70* | IIIB | F | 29 | CS | 90 | 22 | PanC | TC | Central | No | Yes | Yes |
| *sample_007* | m | *25-50* | IIIA | | | | 95 | | PanC | TC | Central | Focal | Yes | No |
| *sample_008* | f | *25-50* | IIA | F | 45 | NS | 80 | 18 | PanC | TC | Central | No | No | No |
| *sample_009* | f | *>71* | IA | T | 41 | NS | | 26 | NeU | AC | Peripheral | Prevalent | Yes | Yes |
| *sample_010* | m | *51-70* | IA | F | 79 | NS | | 31 | NeU | TC | Peripheral | Prevalent | No | No |
| *sample_011* | f | *>71* | IA | F | 63 | ES | 90 | | NeU | TC | Peripheral | Focal | No | No |
| *sample_012* | f | *>71* | IIIA | T | 39 | NS | 80 | 30 | NeU | AC | Central | Focal | Yes | No |
| *sample_013* | f | *51-70* | IIIA | F | 77 | ES | | 30 | NeU | TC | Central | Focal | Yes | No |
| *sample_014* | f | *51-70* | IA3 | F | 43 | NS | 90 | 22 | NeU | AC | Peripheral | Focal | Yes | Yes |
| *sample_015* | f | *>71* | IIB | F | 47 | NS | 90 | | NeU | TC | Peripheral | Focal | Yes | No |

Abbreviations: Gender: f (female), m (male); Deceased: T (true), F (false); Smoking category: NS (never smoker), EX (ex-smoker), CS (current smoker);

Histology: TC (Typical Carcinoid; AC (Atypical Carcinoid); blank indicates data not available.
